# Development of in-house, indirect ELISAs for the detection of SARS-CoV-2 spike protein-associated serology in COVID-19 patients in Panama

**DOI:** 10.1101/2021.04.30.21256406

**Authors:** Carolina de la Guardia, Giselle Rangel, Alcibiades Villarreal, Amador Goodridge, Patricia L Fernández, Ricardo Lleonart

**Author notes:** equal contributions (Carolina de la Guardia, Giselle Rangel).

## Abstract

COVID-19 is the name of the acute respiratory disease caused by the new coronavirus SARS-CoV-2, a close relative of those that caused the severe outbreaks of SARS and MERS several years ago. Since first appearance on December of 2019, the COVID-19 pandemic has cause extremely high levels of mortality, morbidity, global economic breakdown and the consequent human suffering. While vaccination efforts are not extensive and rapid enough, the main tools to keep the virus under control are still keeping physical distancing, reinforce personal hygiene measures, using masks and early diagnosis of virus infected persons, either symptomatic or not. The main diagnostic test for the confirmation of symptomatic individuals is the detection of viral RNA by reverse transcriptase – quantitative real time PCR (RT-PCR). Additionally, serology techniques, such as ELISA are extremely useful to measure the antibodies generated in humans after virus contact, as well as the direct presence of viral antigens. In this study we aim to assemble and evaluate four ELISAs assays to measure the presence of IgG or IgM specific for the viral Spike protein in COVID-19 patients, using either the full recombinant SARS-CoV-2 Spike protein or the fragment corresponding to the receptor binding domain. As a control, we also analyzed a group of prepandemic serum samples obtained before 2017.

Strong reactivity was observed against both antigens. A few prepandemic samples displayed high OD values, suggesting the possibility of some crossreactivity. All four assays show very good repeatability, both intra- and inter-assay; however, no clear relationship could be detected between positivity and time of sample collection for serology. Receiver operating characteristic analysis allowed the definition of cutoffs and evaluation of performance for each ELISA by estimation of the area under the curve. This performance parameter was high for all tests (AUC range: 0.98-0.995). Multiple comparisons between tests revealed no significant difference between each other (*P* values: 0.24-0.95). Our results show that both antigens are very effective to reveal both specific IgG and IgM antibodies, with high sensitivity (range 0.929-0.99) and specificity (range 0.933-0.977). The estimated congruence with the RT-PCR test, as estimated by Cohen’s Kappa, indicates a high agreement in all cases (range 0.874-0.937). This test will allow health authorities to have a new tool to estimate seroprevalence, and to manage and improve the serious sanitary situation created by this virus.

## Introduction

The new coronavirus disease 2019 (COVID-19) is a highly infectious disease caused by the severe acute respiratory syndrome coronavirus 2 (SARS-CoV-2) that has become a world-wide pandemic with huge human and economic losses^1^. Most patients with COVID-19 develop a mild to severe respiratory illness, while others develop minimally- or asymptomatic infection^2^. Several investigations have shown that asymptomatic patients can also spread the disease^3^. Since COVID-19 was declared a global pandemic, the world has suffered more that 130 million confirmed cases and 2.8 million deaths^4^. The Americas have also been strongly affected, with more than 58 million confirmed cases and 1.4 million deceased patients (as of April 11, 2021)^5,6^. The last available situation report for Panama, dated April 6, 2021, shows 356,373 confirmed cases and 6,131 deceased^7^.

Virus specific reverse transcriptase – polymerase chain reaction (RT-PCR) has become the assay of choice to rapidly detect viral RNA sequences in acute infection of either symptomatic subjects or their contacts^8^. For symptomatic individuals, the gold standard for testing is still the RT-PCR on nasopharyngeal swab samples^9,10^. However, RT-PCR is not devoid of important limitations, such as false-negative results^11,12,13,14,15^, variability in accuracy for different types of specimens^10^, hazardous sample collection procedure^16^ and sensitivity issues^11,16^.

Besides RT-PCR, the use of serological techniques has also become an important tool for the management of the COVID-19 pandemic. In symptomatic individuals, serology testing may also help understand important points, such as the duration of virus specific antibodies^17^. For asymptomatic individuals, serology testing contributes to answer epidemiological questions, including virus exposure in general population or in particular high-risk groups, to plan public health interventions, and to monitor vaccine applications and performance^17^. In fact, we recently evaluated the performance of a rapid lateral flow immunoassay to detect IgM/IgG antibodies against SARS-CoV-2^18^. However, this type of rapid tests may present some limitations, such as not allowing quantitation of antibody titers or suboptimal sensitivity^19^. In contrast, better results may be obtained by tests based on the enzyme-linked immunosorbent assay (ELISA), which are easy and convenient to implement^20^.

The humoral response of humans to the SARS-CoV-2 is apparently dominated by the production of antibodies specific for several structural proteins of the virion, mainly the Spike (S) and the Nucleocapsid (N) proteins^21^. Here we report the use of recombinant S protein, either as a full-length antigen, or just the fragment corresponding to the receptor binding domain (RBD) to assemble in-house, indirect enzyme-linked immunosorbent assays (ELISAs) for the detection of SARS-CoV-2-specific IgG and IgM antibodies. We also report the performance of the tests using a group of local COVID-19 patients and prepandemic serum samples collected before 2017. Our results confirm the usefulness of these antigens for the ELISA, opening the possibility for conducting new local studies for better knowledge and management of the disease in the country.

## Materials and methods

### Patients and samples

The study population represents a sub cohort of a cross-sectional study conducted between April and December 2020^18^. Clinical serum samples were obtained from 102 hospitalized patients confirmed to be positive for SARS-CoV-2 viral infection by reverse transcriptase polymerase chain reaction (RT-PCR, done by a national reference lab) on nasopharyngeal swab testing. All participants develop moderate clinical symptoms and were treated in public hospitals located in Panama and Colon cities. For the purpose of negative control samples, 45 prepandemic serum samples were used from de-identified, bio-banked sera stored at INDICASAT-AIP. These samples correspond to research projects on tuberculosis and serosurveillance, and included 15 from patients with active pulmonary tuberculosis, 15 from patients with latent TB infections and 15 healthy individuals from blood bank. All blood samples were collected following regular procedures and centrifuged at 2000 x g for 10 min to obtain serum specimens. Serum samples were aliquoted and frozen at −80°C until testing. Prior to analysis, all serum samples were heat-inactivated at 56°C for one hour.

### Ethics statement

Clinical serum samples from COVID-19 patients were collected as part of a cross-sectional study registered with the Panama Ministry of Health (No.1462) and was approved by the National Research Bioethics Committee (CNBI; No. EC-CNBI-2020-03-43). Prepandemic serum samples were taken in Colón province, with ethical clearance No. 1131/CNBI/ICGES/11 and No. 125/CBI/ICGES/14. All the study participants were enrolled after informed consent was given.

### Antigens

Recombinant antigens used for ELISA were donated by Dr Florian Krammer, and consisted of 1) recombinant RBD fragment, corresponding to amino acids 319–541 plus a His tail, and 2) the full spike protein, modified as described for stabilization and multimerization (aa 1-1213)^22^. Before use, recombinant protein batches were checked for concentration and integrity by SDS-PAGE (data not shown).

### ELISAs

The ELISA assays were implemented to detect SARS-CoV-2-specific IgG and IgM antibodies specific to S and RBD antigens. Protocol was adapted from Stadlbauer et al (2020)^23^. Briefly, 96-well high binding microtiter plates (Corning Costar no. 3361) were coated with 50 µL of RBD or S recombinant protein (both at 2 µg/mL) in 1X Phosphate Buffered Saline (PBS), overnight at 4°C. The next day, wells were washed three times with 250 µL of PBST (PBS containing 0.1% Tween 20) and blocked for 2 h at room temperature (RT) with 200 µL of 1.5% non-fat dry milk (NFM) in PBST. Plates were then washed as before and serum samples added in 100 µL, prediluted 1:50 in PBST with 0.5% NFM, and incubated 2 h at RT. After similar washing, secondary conjugated antibody was added at 1:3,000 dilution in PBST with 0.5% NFM and incubated for 1 h at RT. Specific IgG and IgM antibodies were detected with anti-human HRP conjugates: goat anti-human IgG (H+L), Thermo Fisher No. H10307, or goat anti-human IgM (Heavy chain), Thermo Fisher No. A18835, respectively. Plates were then washed and 100 µL of substrate TMB (3, 3’, 5, 5’ – Tetramethylbenzidine Liquid substrate, Super Sensitive, Sigma) added and incubated for 5 min. Reaction was stopped with 50 µL of 1M HCl and immediately read at 450 nm (SynergyHT, BioTek).

### Statistical analysis

All statistical analyses were carried out using Prism v9.1.0 (GraphPad, USA) and easyROC online calculator (http://www.biosoft.hacettepe.edu.tr/easyROC/). Results of data analysis are presented as estimated statistics and their 95% confidence intervals (CI), or coefficients of variation (%CV). Normality of the optical density data was tested using Kolmogorov-Smirnov procedure. Correlation between OD data and evolution time (time elapsed between the positive RT-PCR result and serum sampling) was assessed using non parametric Spearman’s rank-order correlation test. Receiver-operating characteristic (ROC) analysis was performed on ELISA OD data to calculate area under the curve (AUC) and optimal cut-off, based on procedures implemented in online calculator easyROC^24,25^. Pairwise comparisons between the AUCs of described ELISA tests were done using the multiple comparison procedure implemented in easyROC. The performance of the tests was assessed by calculating sensitivity, specificity, and positive- and negative predictive values, associated *P* values and their 95% confidence intervals using Fisher exact test as implemented in GraphPad Prism. Based on cutoff calculated in the ROC analysis for each test, samples were classified as positive or negative, and agreement between RT-PCR test and ELISAs was assessed using Cohen`s kappa statistic, calculated as described by Fleiss et al 2003^26^, using an online calculator (GraphPad QuickCalcs, USA). For all statistical analyses, *P* values < 0.05 were considered statistically significant.

Repeatability assessment was done by selecting two positive serum samples, including one strongly positive (OD 2) and one weakly reactive (OD 0.6), and one negative. With these samples, ELISAs were repeated, in quadruplicates, over at least five different days. Then, coefficients of variation were calculated both intra- and inter-assay using OD data.

## Results

We assembled and evaluated in-house, indirect ELISAs to detect SARS-CoV-2 specific IgG or IgM antibodies in serum samples, using two recombinant viral proteins, the receptor binding domain of spike (RBD), or the full-length trimeric spike as previously reported^22^. For this study, we used serum samples from two groups, 102 COVID-19 patients and 45 prepandemic serums obtained from patients enrolled in previous studies (**Table 1**). COVID-19 patients were enrolled from May 4 to December 27, 2020 at two hospital wards including: Complejo Hospitalario Dr. Arnulfo Arias Madrid in Panama City and Hospital Manuel Amador Guerrero, Colon City. All these patients had moderate symptoms and a positive RT-PCR test (median time of RT-PCR to serology, 14 days). Main clinical signs at admission were fever (76%), dysnea (72%) and cough (68%). Additionally, 73% reported to had at least one chronic disease, including hypertension (47%), type II diabetes (36%), and renal failure (11%) (data not shown). Prepandemic samples were obtained from subjects participating in tuberculosis (TB) research and surveillance programs at the Colon province, Panama.

**Table 1.**
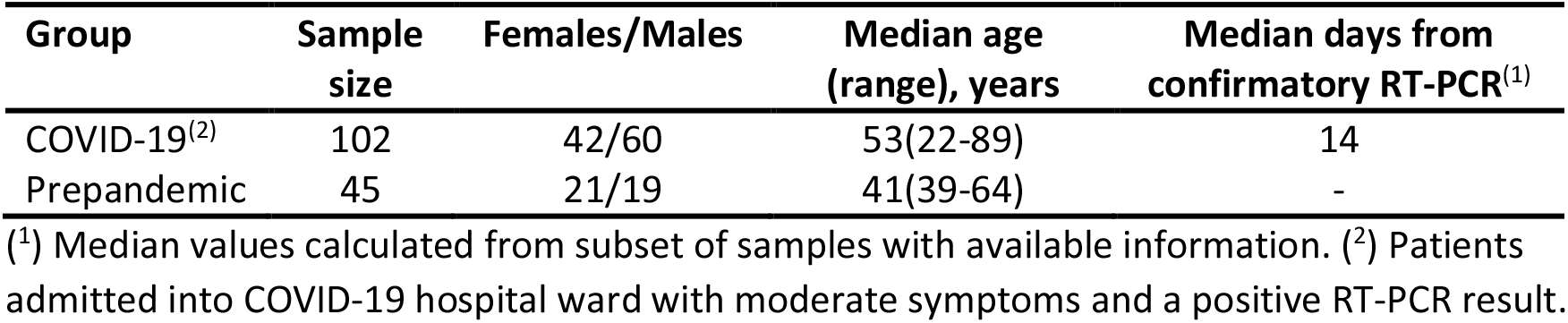
Summary of samples used in this study.

Some COVID-19 samples were negative for all ELISA tests assayed, irrespective of their time of sampling after the RT-PCR positive result (data not shown). These particular samples also tested negative previously in a rapid immunochromatography serology test measuring IgG and IgM ^18^, therefore were considered non-responsive and not considered for the rest of the study. When using 1:50 dilution of samples, strong reactivity was observed in most COVID-19 samples, while vast majority of prepandemic ones remain negatives (**Figure 1**). Some overlapping of OD values between COVID-19 and prepandemic samples was observed for all assays. IgG detecting tests showed a more concentration of OD values in a strongly positive or negative zone, while OD values produced by IgMs were less defined regardless of the antigen used. A few samples (5) in the prepandemic group showed higher than expected values at some of the tests, particularly in the RBD-IgG ELISA (**Figure 1**), suggesting potential crossreactivity with pre-existing antibodies.

**Figure 1.**
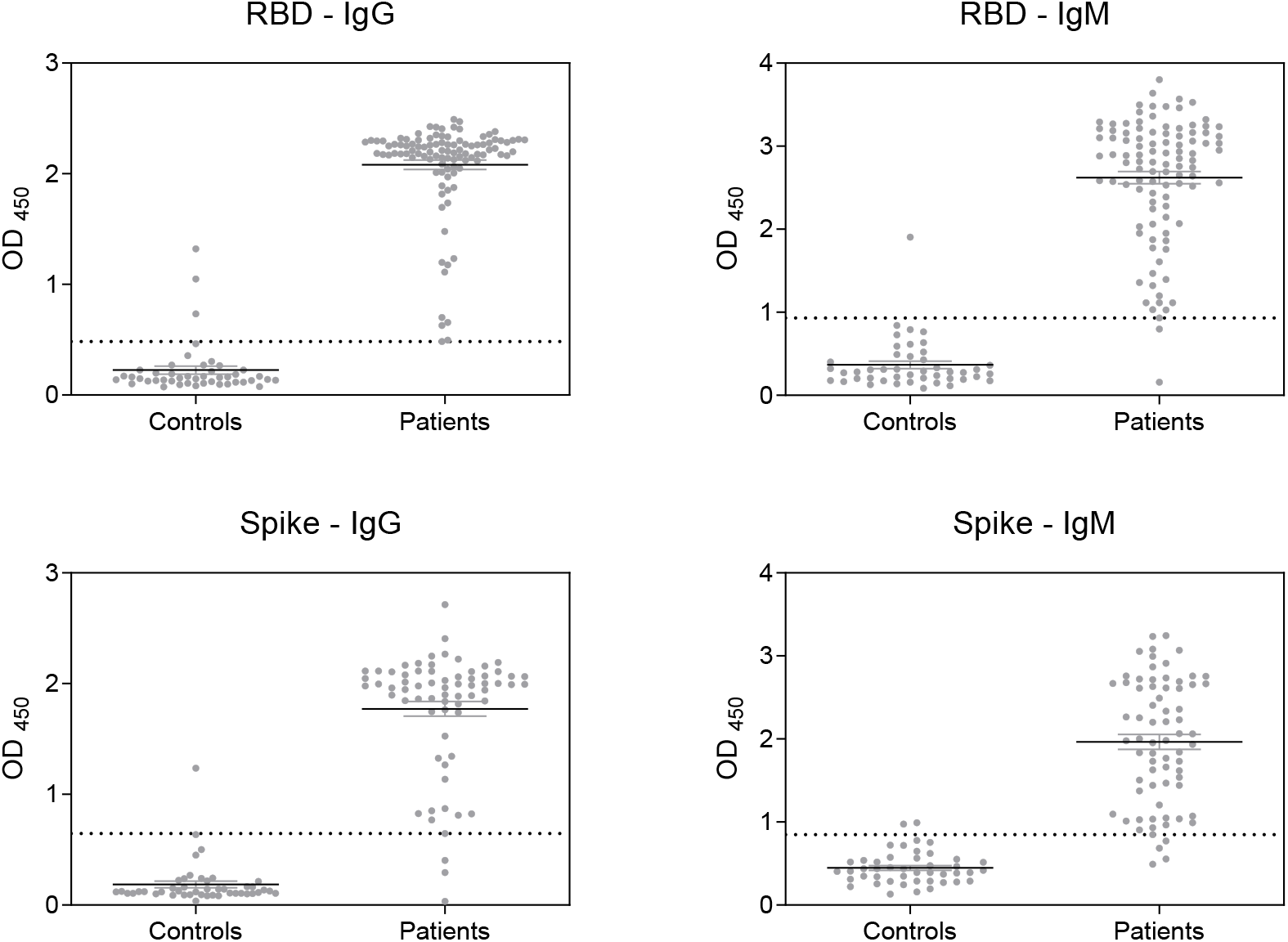
Serum reactivity of COVID-19 patients and prepandemic controls at each ELISA. RBD: recombinant spike receptor binding domain. Spike: recombinant full Spike protein. Samples were processed as described in Materials and Methods. Dotted lines show the optimal cutoff for each ELISA test as determined by ROC analysis (Table 2).

For further evaluation of the performance of each ELISA, receiver-operating characteristic (ROC) analysis was performed using OD data (**Table 2**) and the area under the curve (AUC) graphs were constructed to graphically depict performance of the systems (**Figure 2**). The area under the curve was high for all ELISA systems, with highest value for the RBD-IgG ELISA (range 0.98-0.995) suggesting an excellent performance in all cases. By performing all possible pairwise multiple comparisons of AUC for all ELISA tests, we could not observe significant differences, with *P* values ranging from 0.24 (RBD-IgG vs Spike-IgM) to 0.95 (RBD-IgM vs Spike-IgM), suggesting a very high performance yet very similar for all tests. Using Youden approach for selecting cutoffs based on both sensitivity and specificity, the best values were estimated in a range of 0.48 to 0.93 units of absorbance (**Table 2**). Using these calculated cut-off values, samples were classified as positive and negative for subsequent analyses.

**Table 2.** Performance of ELISA tests as estimated by ROC analysis.

| Performance measures | ELISA assay |  |  |  |
| --- | --- | --- | --- | --- |
|  | RBD<sup>(1)</sup>-IgG | RBD-IgM | Spike<sup>(2)</sup>-IgG | Spike-IgM |
| AUC<sup>(3)</sup> (95% CI) | 0.995 (0.99-1) | 0.987 (0.969-1) | 0.98 (0.95-1) | 0.986 (0.972-1) |
| AUC standard error | 0.002 | 0.009 | 0.015 | 0.007 |
| Optimal cut-off<sup>(4)</sup> | 0.483 | 0.933 | 0.646 | 0.847 |
| Sensitivity (95% CI) | 0.990 (0.946-0.999) | 0.970 (0.917-0.992) | 0.941 (0.858-0.976) | 0.929 (0.845-0.969) |
| Specificity (95% CI) | 0.933 (0.821-0.977) | 0.977 (0.884-0.998) | 0.977 (0.884-0.998) | 0.955 (0.851-0.992) |
| PPV<sup>(5)</sup> (95% CI) | 0.971 (0.918-0.992) | 0.990 (0.945-0.999) | 0.984 (0.917-0.999) | 0.970 (0.899-0.994) |
| NPV<sup>(6)</sup> (95% CI) | 0.976 (0.879-0.998) | 0.936 (0.828-0.978) | 0.916 (0.804-0.967) | 0.895 (0.778-0.954) |
| Kappa (<sup>7</sup>) (95% CI) | 0.935 (0.873-0.998) | 0.937 (0.876-0.998) | 0.909 (0.831-0.987) | 0.874 (0.784-0.964) |
| Kappa SE | 0.032 | 0.031 | 0.040 | 0.046 |

(<sup>1</sup>) RBD, recombinant SARS-CoV-2 Receptor Binding Domain. (<sup>2</sup>) Spike, recombinant SARS-CoV-2 Spike protein. (<sup>3</sup>) AUC, area under the ROC curve. (<sup>4</sup>) Calculated by Youden method. (<sup>5</sup>) Positive Predictive Value. (<sup>6</sup>) Negative Predictive Value. (<sup>7</sup>) Agreement with RT-PCR assay.

**Figure 2.**
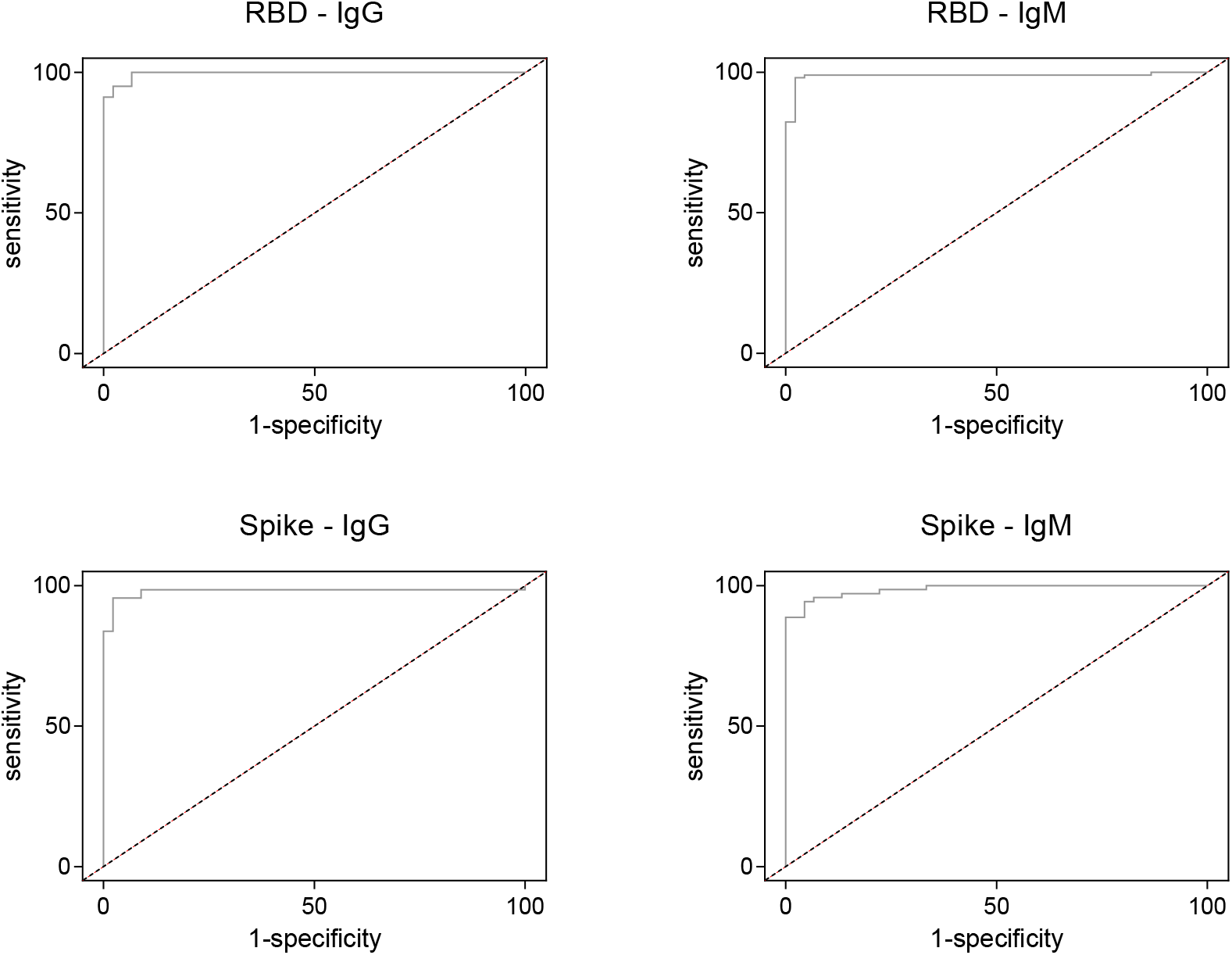
Performance of the described ELISA tests by receiver-operating characteristic (ROC) analysis. RBD: recombinant spike receptor binding domain. Spike: recombinant full Spike protein. Dotted lines represent the theoretical performance of a test with no discriminatory ability, corresponding to an area under the curve (AUC) of 0.5.

All four assays showed good estimated values for sensitivity, specificity, positive and negative predicted values, and Kappa statistics. Sensitivity values were higher for the RBD-IgG ELISA (0.99, range 0.929-0.99), while specificity was better for the RBD-IgM and Spike-IgG ELISAs (both 0.977, range 0.933-0.977). Positive predictive values showed highest results for RBD-IgM (0.99, range 0.97-0.99), while negative predictive value was best for the RBD-IgG ELISA (0.976, range 0.895-0.976). Although the RT-PCR and ELISA tests would probably have very different purposes, strengths and performances, we also measure the agreement by estimating Kappa statistic. The calculated values of Cohen`s kappa (range 0.874 – 0.937) indicated an “almost perfect agreement” between both types of assay (**Table 2**), as per the interpretation scale of Landis and Koch (1977)^27^.

We attempted to evaluate if there was a relationship between the time of evolution (time after the positive RT-PCR result) and optical density data for each ELISA. Normality of the OD data sets was tested using Kolmogorov-Smirnov test, resulting in rejection of normality. Therefore, correlation between OD and time variables was assessed using non parametric Spearman’s rank-order correlation test. Optical densities did not show significant correlation with evolution time for any of the described ELISAs (RBD-IgG: *P*= 0.19; RBD-IgM: *P*=0.80; Spike-IgG: *P*=0.13; Spike-IgM: *P*= 0.43; **Supplementary Figure 1**). The same was observed by looking at the positivity rate for time period slots (**Table 3**).

**Table 3.** Positivity rate by time in COVID-19 group.

| Time (days)<sup>(1)</sup> | Positivity rate for each ELISA assay |  |  |  |
| --- | --- | --- | --- | --- |
|  | RBD-IgG | RBD-IgM | Spike-IgG | Spike-IgM |
| 0-7 | 7/7 | 7/7 | 5/6 | 7/7 |
| 8-14 | 31/32 | 31/32 | 15/16 | 15/17 |
| 15-21 | 22/22 | 22/22 | 5/5 | 6/7 |
| >22 | 16/16 | 14/16 | 14/15 | 14/15 |
| Total | 76/77 | 74/77 | 39/42 | 42/46 |

(<sup>1</sup>) Time after RT-PCR confirmation.

To assess the repeatability of the assays we also measure the coefficient of variation (%CV) both intra- and inter-assays, by selecting several negative, weak positive and strong positive sera and measure the dispersion of the resulting ODs at several replicates within a plate and among several plates in different days (**Table 4**). The values of CV ranged from 4% to 9%, well below the maximum tolerated of 15% for ELISA tests, indicating very good performance in terms of repeatability.

**Table 4.**
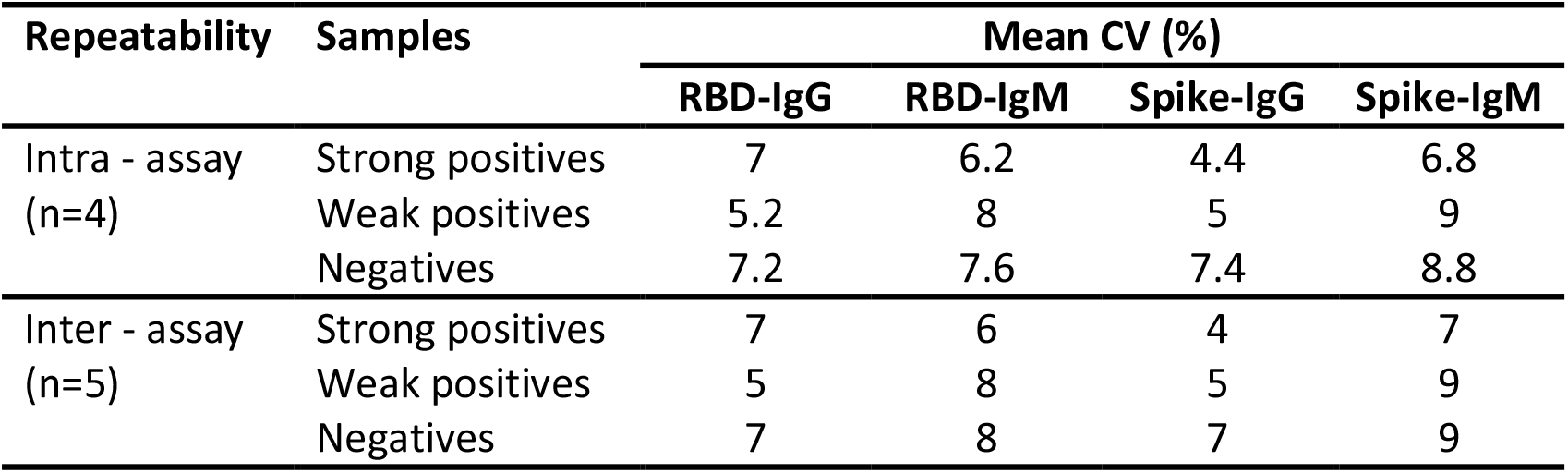
Repeatability of the described ELISAs, based on calculated intra-assay and inter-assay coefficients of variation.

| Repeatability | Samples | Mean CV (%) |  |  |  |
| --- | --- | --- | --- | --- | --- |
|  |  | RBD-IgG | RBD-IgM | Spike-IgG | Spike-IgM |
| Intra - assay<br>(n=4) | Strong positives | 7 | 6.2 | 4.4 | 6.8 |
|  | Weak positives | 5.2 | 8 | 5 | 9 |
|  | Negatives | 7.2 | 7.6 | 7.4 | 8.8 |
| Inter - assay<br>(n=5) | Strong positives | 7 | 6 | 4 | 7 |
|  | Weak positives | 5 | 8 | 5 | 9 |
|  | Negatives | 7 | 8 | 7 | 9 |

## Discussion

In this study, we have evaluated four in-house ELISAs for the detection of antibodies against SARS-CoV-2 proteins. RBD- and full S-based ELISAs were used to detect IgM and IgG antibodies in serum samples of RT-PCR confirmed COVID-19 patients and pre-pandemic samples. As expected, most COVID-19 patients showed a strong reactivity of both types of antibodies against both the full Spike protein and the receptor binding domain. As well, most of the prepandemic serum samples remains unreactive. However, some overlap was observed in the OD values of both groups of samples, for all four assays. Some samples in the COVID-19 group remain negative, a fact already observed by some authors that suggests a lack of- or a very weak seroconversion^28,29,30^.

In terms of the temporal behavior of the seroconversion, we did not observe a clear pattern of increase positivity in time. In fact, most of our COVID-19 group samples show very early seroconversion, while some remain negative irrespective of their time since RT-PCR confirmation. This unusual trend is consistent with other reports indicating a much earlier seroconversion in COVID-19 patients^31,32,33^ and a non-typical temporal pattern of responses for IgG and IgM^34^.

Interestingly, some of the samples of the pre-pandemic group showed high levels of reactivity, particularly in the RBD-IgG ELISA, suggesting possible crossreactivity with pre-existing antibodies. Several authors have shown antibody crossreactivity in the immune response to several coronaviruses in human samples^28,35,36,37^.

Apart from highly pathogenic SARS-CoV, MERS-CoV and SARS-CoV-2, infections by other human coronaviruses are seldom reported, particularly in Panama and other countries of the region^38^. These viruses, however, are also able to cause severe disease in children, elderly patients and immunocompromised individuals^39^. Although heavily underreported, these coronaviruses have been circulating in several countries of the region, with estimated prevalence of about 5-7% of all influenza-like infections in Latin American countries including Brazil, Costa Rica and Colombia^40^. Although we could not assess the contact status of our prepandemic samples with other human coronaviruses, it is conceivable that some of them may have been exposed to these endemic coronaviruses and potentially develop crossreactive antibodies.

Our study presents however some limitations. The number of samples analyzed is still low. Additionally, to assess the relationship between positivity and evolution time we used the available date of RT-PCR confirmation, and this may not be a good surrogate of infection time. When available, studies should try to use the date of symptoms onset for better estimation of seroconversion time. It is also important to note that our COVID-19 cohort only include mild to moderate patients, and this fact may have influenced the strength of the immune response observed in our study. It has been shown that severity correlates with a more intense immune response and production of specific antibodies^41,42,43,44,45^. A more complete picture of the serology would have been obtained with patients from all the spectrum of the disease. Next steps should include testing of RT-PCR positive, asymptomatic subjects to characterize whether the tests are able to detect lower antibody levels likely expected in such situation.

Nevertheless, our results confirm previous reports indicating the usefulness of these antigens for assays to measure seroconversion after SARS-CoV-2 infections^22,46,47,48^. The ELISA tests are simple to perform and very robust once optimized. Besides, since only a very small amount of sample is required, they are very convenient. Our results are consistent with those shown by others, suggesting that these serology assays have a very good potential for studies about measuring virus exposure in large groups, as well as seroconversion and seroprevalence studies in Panama. Additionally, since current vaccines being applied in Panama are based on Spike protein as immunogen, these assays may be a useful asset to study their effect and monitor the general immunological status of the population on the way to the much-needed herd immunity.

## Data Availability

Data not shown in the manuscript are available from authors upon reasonable request.

## Acknowledgements

We are very thankful to Dr. Florian Krammer (Department of Microbiology, Icahn School of Medicine at Mount Sinai, New York, NY, USA) for donation of the SARS-CoV-2 recombinant proteins used in this study. We thank all enrolled patients for their willingness to participate in the study. We are very grateful to Dr. Eduardo Ortega and Dr. Jagannatha Rao for continued support and useful advice during the course of this study. We also thank MSc. Dilcia Sambrano, Dr. Carlos Restrepo, Dr. Diana Oviedo, BSc. Ambar Pérez-Lao, Dr. Odemaris Luque, Dr. Ana de Chavez, Dr. Julio Jurado, Dr. Nicolas Hurtado, Dr. Azael Gonzalez and BSc. Laiss Mudarra at Caja de Seguro Social and Ministerio de Salud in Colon City for their support during recruitment and sampling. We also thank support of Sistema Nacional de Investigación (SNI, SENACYT, Panama) for the authors A.V., A.G., P.L., and R.L.

## Authors contributions

A.V., A.G., P.L. and R.L. conceived of the idea and design the experiments. C.G. and G.R. perform assay set up and antibody determinations. G.R. and R.L. analyzed the data. P.L. and R.L. wrote the paper with input from all authors.

## Tables and figures

**Supplementary Figure 1.**
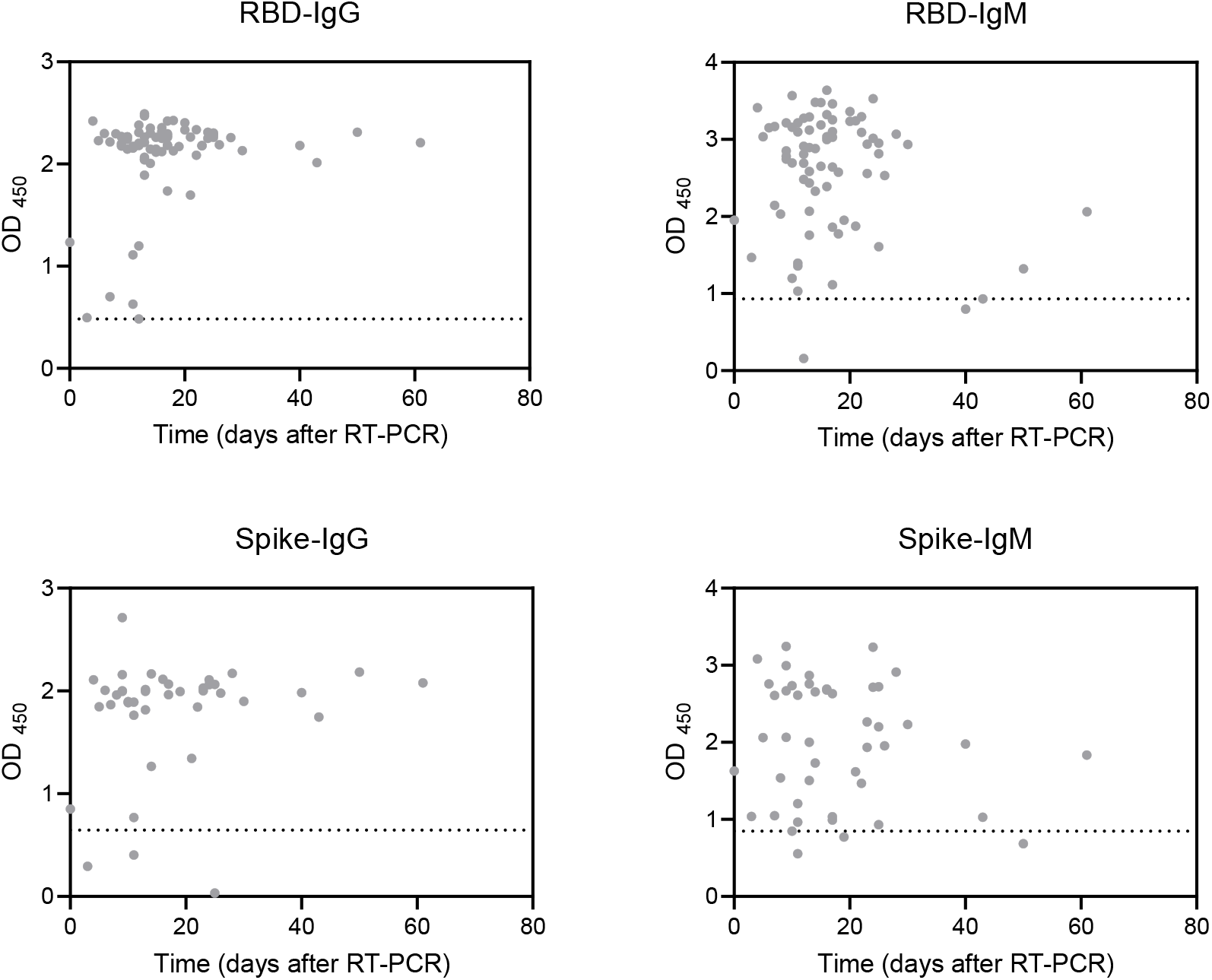
Scatter plots showing the reactivity of each sample in ELISA as optical density at 450 nm, with their corresponding sampling time (since the positive RT-PCR result), for those samples for which data was available. RBD: recombinant spike receptor binding domain. Spike: recombinant full Spike protein. Dotted lines represent the cutoff value for each ELISA.

